# Feasibility of Reusing Surgical Mask Under Different Disinfection Treatments

**DOI:** 10.1101/2020.05.16.20102178

**Authors:** C.Y. Suen, H.H. Leung, K.W. Lam, K.P.S. Hung, M.Y. Chan, Joseph K.C. Kwan

## Abstract

The possibility to extend the lifespan or even reuse one-off personal protective equipment, especially for N95 respirator and surgical mask become critical during pandemic. World Health Organization has confirmed that wearing surgical mask is effective in controlling the spread of respiratory diseases in the community, but the supply may not be able to satisfy all the demands created all over the world in a short period of time. This investigation found that dry heat and UVC irradiance could effectively disinfect the mask material without creating significant damage to surgical mask.

## I. Introduction

During pandemic, the huge demand of personal protective equipment (PPE) compels countries to investigate the possibility to reuse and extend the lifespan of every one-off PPE, especially for N95 respirator and surgical mask.

World Health Organization (WHO) has confirmed that wearing surgical mask is effective in controlling the spread of certain respiratory viral diseases in the community [1]. However, the supply of surgical masks is still behind the huge demand created all over the world. In order to provide an alternative for the general public to maintain basic protection with limited resource, the possibility of reusing surgical mask after different disinfection methods was investigated. Methods of disinfection studied include 100° dry heat, steaming, boiling, autoclave, 75% and 95% ethanol, UVC irradiance and household detergent.

All the disinfection treatments used for disinfecting surgical mask were proven to be effective [2], except household detergent. However, studies of disinfection efficiency, structural and property changes of surgical mask after various disinfection treatments were very limited.

ASTM-F, EN14683, KF and YY0469 are common standards for testing the quality of surgical masks. Although the testing parameters are not completely the same among these standards, they all involve testing of filtration efficiency, breathability and fluid repellency. Filtration efficiency typically considers the effectiveness of particles of aerodynamic size from 0.1µm to 3µm. The cut-off sizes used for the standard tests are classified as particulate filtration efficiency (PFE) and bacteria filtration efficiency (BFE) [3]. Latex sphere is used for the PFE filtration test with the ASTM standard test [4] while the KF, YY0469 and NIOSH use sodium chloride aerosol to perform standard tests for respirator and surgical mask [5]–[7]. Since droplets will start evaporating when exposed to air, the terminal velocity decreases when the droplet size reduces [8]. This implies that small-sized droplets can travel a longer distance, resulting in a higher possibility of infection if the droplet nuclei is a pathogen.

In assessing the destructive level of different disinfection treatments to surgical masks, filtration efficiency and fluid repellency were the parameters being focused in this study. Structural changes of filtration layer were observed and verified after the treatments. The effectiveness of disinfection to surgical mask was thoroughly investigated.

## II. MATERIALS AND METHOD

### Mask selection and disinfection treatment methods

Medicom ASTM level 1 surgical mask was chosen as the test subject due to its popularization. This surgical mask model is widely used in hospitals and health care sectors. The samples were labelled with numbers and randomly assigned to various treatment groups. Each treatment group (N=3) was disinfected by one of the following methods:

a. Dry heat – An oven (Breville BOV820BSS, 2400 W) was used for heating at 100° for 15 minutes. The temperature was monitored by the LCD display of the oven with reference to an infrared thermometer.
b. Steaming – Samples were placed at the centre of a steamer cooker at 100° for 10 minutes.
c. Boiling – Samples were placed at 100° water bath for 10 minutes.
d. Autoclave – Samples were wrapped with aluminum foil individually and placed into the middle basket of an autoclave (Hirayama, HVE-50). The autoclave sterilization was set at 121° for 20 minutes (complete cycle time was 1.5 hour).
e. Detergent – 0.5% w/v of household detergent (Ultra Axion) was prepared by dilution with DI water. Samples were submerged into the solution for 30 minutes, and then gently rinsed with DI water for 1 min to wash away the remaining detergent.
f. UVC irradiation – Irradiation disinfection was performed by using a biosafety cabinet (NUAIRS, NU-425-400S) fitted with 254 nm (20W) tube for 10 minutes. Power density at the mask level was monitored with UVX Radiometer with UVX- 25 254nm UV probe. Samples were placed at a distance of 60 cm from the tube for irradiation with measured average UVC intensity of 450µW/cm^2^. The samples were turned over at 5 minutes for even irradiation.
g. Samples were submerged into 95 % v/v ethanol for 5 minutes.
h. Samples were submerged into 75 % v/v ethanol for 5 minutes.

All samples were air dried in a biosafety cabinet for 24 hours before all measurements.

### Filtration efficiency test

2% sodium chloride solution (NaCl) was aerosolized by Particle Generator Model 8026 with a count median diameter at 0.04 micrometer (nominal) and a geometric standard deviation of 2.2 (nominal). DUSTTRAK™ II Aerosol Monitor Model 8532 was used to measure the particle concentration before and after filter material. An impactor for 1µm was installed to the Aerosol Monitor to study the penetration of small droplets. Each sample was fitted to the sample holder and fastened by screws and rubber O-rings. The penetration of sample was measured when instruments were operating after 2 minutes for stabilization.

Measurements were taken at the sampling holes perpendicular to where *n_f_* is bactericidal efficiency the laminar flow at aerosol upstream and downstream of the sample. The measurement of the concentration of NaCl droplet was taken at a face velocity of 14 cm/s. The filtration efficiency was calculated by the following equation:

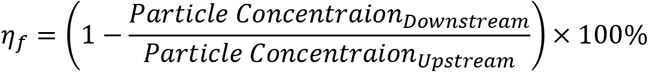

where *η_f_* is Filtration efficiency

(The background particle count was cancelled before calculation.)

**Fig. 1.**
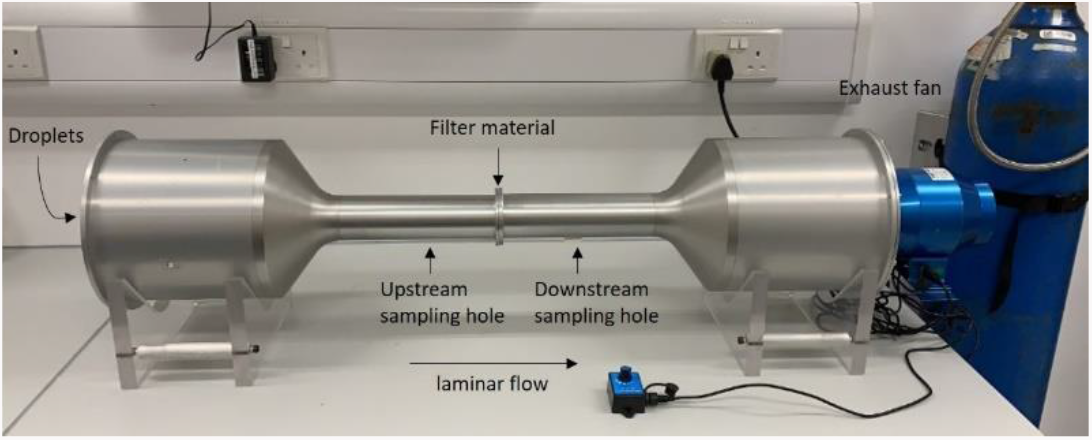
Setup for filtration efficiency test

### Bactericidal test

*Staphylococcus aureus* (clinical isolate) was inoculated to nutrient broth no.2 (Thermo Scientific^™^ Oxoid^™^ Nutrient Broth No.2 (Dehydrated)) by an inoculating loop aseptically. The bacteria were then cultivated overnight in an environmental shaker at 37°C, 200 rpm. The overnight culture would be re-cultured for a few hours until it reached the concentration of 10^8^ CFU/mL. Bacteria culture prepared was diluted to 10^6^ CFU/mL with 0.9% saline and 0.1% tween 80. Mask was cut into 1 inch x 1 inch square and put on supporting glass plate of 1 inch x 1 inch square with external layer of the mask material facing upward.

50 µL of the diluted bacteria suspension was dropped onto each sample and the inoculum was spread evenly to allow soaking into the sample. Three samples were picked randomly and treated with the aforementioned disinfection methods.

A timer was used to monitor the exposure time of the samples with bacteria. Afterwards, the samples with supporting glass plates were transferred to a sterile bottle containing 10 mL of extraction solution (0.9% saline, 0.1% Tween 80). The bottle was vortexed for 20 seconds, allowing sufficient time for dislodgement of microbes into the solution. The suspension was serially diluted with sterilized 0.9% saline solution. 100 µL of the solution was inoculated into TSA agar and cultured at 37°C for 24 hours. The Colony Forming Units (CFUs) on agar plates were enumerated. The Bactericidal Efficiency was calculated by the following equation:

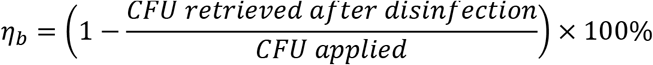

where *η_b_* is bactericidal efficiency

### Hydrophobicity test

Five 100uL load of DI water were added at a distance of 10 cm above the sample at 5 different spots on the outer layer of mask. Visual Inspection of droplets was carried out for each sample. After wobbling the mask gently, hydrophobicity was recorded if the water droplets hold as beads. Hydrophilicity was recorded when water droplet shape was flattened (lowered contact angle) but did not penetrate all three layers. Thorough damage of the water repelling layer was recorded when the water droplet was absorbed and penetrated to the bottom.

### Structural deformation investigation

The filtering layer (polypropylene) of each sample after treatments were cut into a size of 4 mm x 4 mm and fixed on copper stage with carbon tapes. The samples were observed with a scanning electron microscope (SEM) (Hitachi, TM3000) at magnification of 1000×. Structural changes such as melting, deformation, entanglement or cracking of polypropylene fibre were recorded.

### Data Analysis

Comparison of the control and treatment groups were analyzed by t-test in SPSS (version 19). P values <0.05 were considered significant.

## III. Results

All eight disinfection methods did not cause observable change to the samples, except for the steamed and boiled samples which demonstrated a tarnished and softened appearance respectively. Earloop of the samples after boiling, steaming and autoclave treatment lost elasticity.

SEM was used to observe any micro structural change of masks after different treatments. SEM images at 1000× revealed that all methods used in the experiment did not cause observable structural change to the filtering layer. No shrinkage, melting, deformation, entanglement or cracking of fibre was noted.

All samples were tested for filtration efficiency with NaCl droplets. Control samples had an average filtration efficiency of 97.83 % ± 0.9 % at 14 cm/s. The average filtration efficiency of household detergent water treated and ethanol treated samples were significantly lower than the control. Samples after boiling, steaming, baking & UVC irradiation were slightly offset from the control but were not significantly different.

Bead-like droplets were observed in all the samples treated with non-fluid disinfection methods, such as dry heat and UVC irradiation. For the samples underwent other disinfection treatments, the fluid-repelling layers were concluded to be damaged as the water droplets on the mask surface could not retain bead shape.

All traditional boiling, steaming, submersion in ethanol and dry heat methods were effective in reducing the *S. aureus* load for 4-log. Irradiation with UVC could also achieve the same extend of disinfection in mask material.

**Fig. 2.**
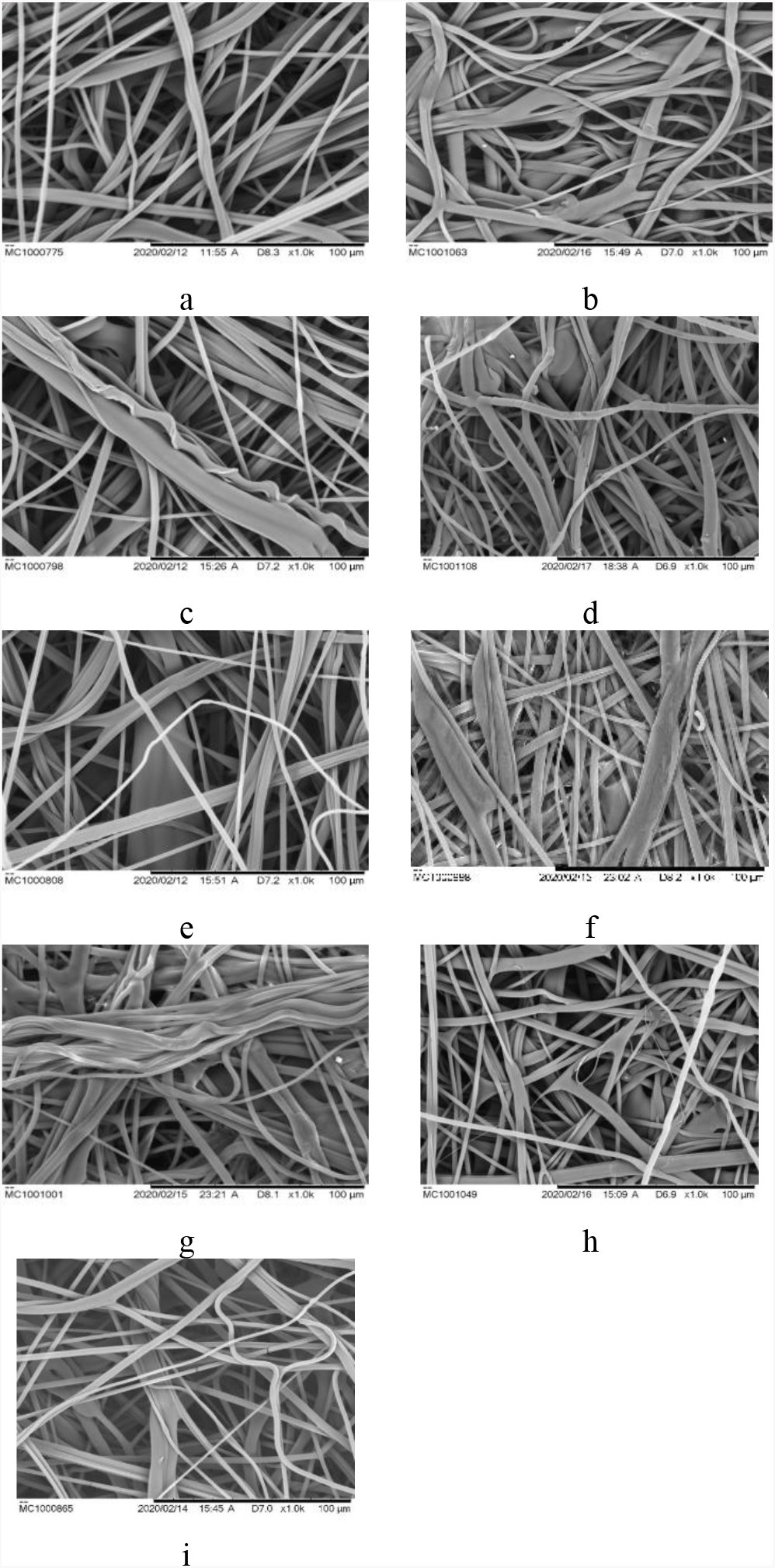
Appearance of filtration layers of masks after different treatments under scanning electron microscope at 1000×. a: control, b: boiled sample, c: steamed sample, d: autoclaved sample, e: detergent treated sample, f: 75% ethanol treated sample, g: 95% ethanol treated sample, h: dry heat sample, i: UVC treated sample

**Fig. 3.**
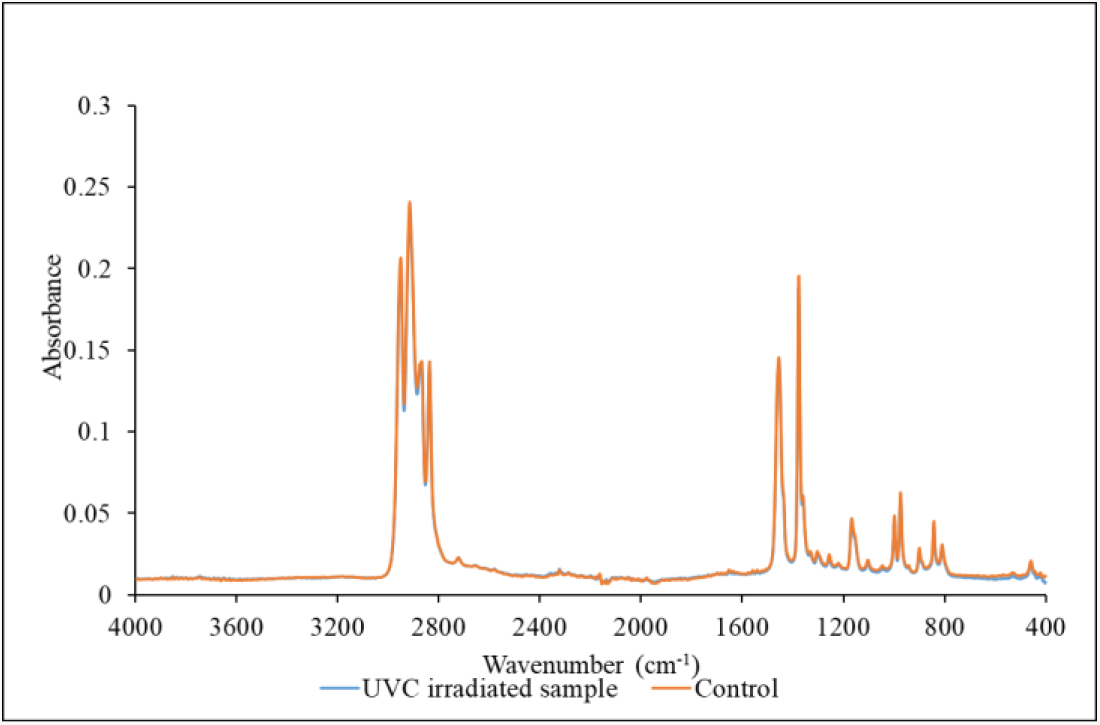
FTIR spectrum of surgical mask (filtration layer). Orange curve indicates the control sample while blue curve indicates the UVC treated sample at 450µW/cm^2^ for 90 minutes.

UVC treated sample was scanned using FTIR. Result showed insignificant difference between control and treated samples.

## IV. Discussion

Due to the pandemic, limited supply and a sudden surge of need for surgical masks in many places have been reported. Surgical mask decontamination methods have been reported from various media. The goal of the study was to evaluate some common surgical mask decontamination methods and to identify the possible methods that cause the least damage to the surgical mask in terms of overall integrity, hydrophobicity and filtration rate. As practical decontamination methods should aim at effectively eliminate harmful microorganisms with high reproducibility while remaining safe to users, certain decontamination methods such as chlorine gas and ozone gas treatment were not included in this experiment.

Certain drawbacks have been noted in some preliminary tests, for example, aqueous 1:99 bleach solution alone cannot penetrate into the mask to perform thorough disinfection. Ozone gas is highly hazardous to human being [9]. It is not recommended to be used in area without proper ventilation. Chlorine gas and other chlorinated compounds were known to be incompatible with polypropylene [10]. Treatment with incompatible disinfectants may cause unwanted damages to the surface characteristics and structure of the mask.

### Disinfection Efficiency

Except household detergent water, all disinfection methods were effective in eliminating *S.aureus* in the mask material. Traditional disinfection methods such as boiling, steaming, submersion in ethanol, dry heat, UVC irradiation have long been used to control microbial growth. *S.aureus* is a gram-positive bacteria commonly used to demonstrate efficiency of various disinfection methods.

Moist-heat treatment methods, including boiling, steaming, autoclave, expose the mask material to 100 % R.H. but not in the case of dry heat treatment. Disinfection by dry heat has been known to be relatively slower than moist-heat, however, dry heat at 100 °C for 15 minutes was still efficient to achieve a 4-log reduction.

Ethanol treatment between 60% to 95% denatures protein and dehydrate microorganisms[2]. Results from the experiment also showed an efficient 4-log reduction of *S.aureus* in 5 minutes.

254 nm UVC irradiation is a physical disinfection method that utilizes electromagnetic wavelength to damage RNA, DNA, and protein. The samples were turned over after 5 minutes because the irradiation may not be efficient to penetrate the entire mask material and eliminate “dead-corner”. Experimental result showed the exposure at 450 µW/cm^2^ for a total of 10 minutes could effectively eliminate all *S. aureus* in the mask material.

Household detergent was not classified as a disinfectant. The detergent used in the experiment did not have bactericidal effect towards *S. aureus*. Reduction demonstrated in table 1 could be a result of the partial suspension of bacteria into the solution.

**Table 1.**
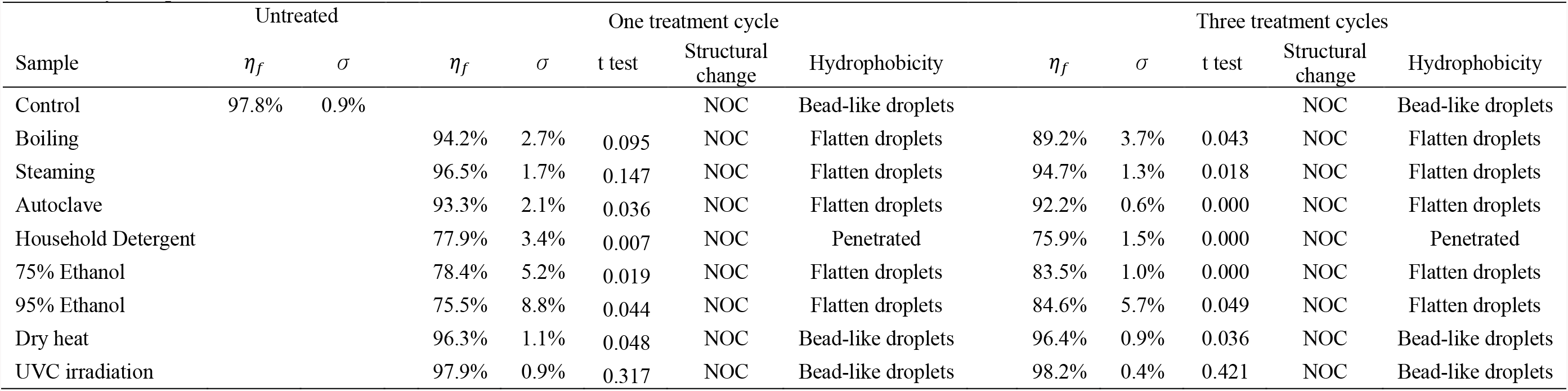
Physical performance

**Table 2.** Disinfection efficiency

| Sample | Reduction | $\sigma$ |
| --- | --- | --- |
| Control | - | - |
| Boiling | 99.99% | ± 0% |
| Steaming | 99.99% | ± 0% |
| Autoclave | 99.99% | ± 0% |
| Household detergent | 50.86% | ± 9.2% |
| 75% Ethanol | 99.99% | ± 0% |
| 95% Ethanol | 99.99% | ± 0% |
| Dry heat | 99.99% | ± 0% |
| UVC irradiation | 99.99% | ± 0% |

### Hydrophobicity test

Dry heat and UVC treatment did not show observable effect on the hydrophobicity of mask surface. For household detergent, rinsing with water might not completely remove detergent from mask material. All detergent treated masks demonstrated severe water penetration possibly be related to the lowered surface tension [11]. For treatment by boiling, steaming, autoclaved and ethanol, samples demonstrated flattened droplets. This could possibly be related to altered surface characteristic after treatments.

### Structural changes

Most treatment groups did not demonstrate observable change of the general structure of surgical mask. All the ear loops held unchanged throughout the experiment, except for the samples treated by boiling, steaming and autoclave. Treatment by boiling and steaming had generally softened the texture of the mask.

UVC did not damage the filtration layer even the exposure time was up to 90 minutes. Although UVC is proven to facilitate the degradation of polypropylene, short exposure targeted for disinfection of masks would not cause significant structural damage to the filtration layer [12].

### Filtration Efficiency

After one treatment cycle, all treatment methods except UVC demonstrated a drop in filtration efficiency. Treatment with household detergent and ethanol resulted in significant decrease of the filtration efficiency. After three treatment cycles, treatment by UVC and dry heat could still maintain high filtration efficiencies (> 95%).

Generally, non-fluid treatments perform better than fluid-based treatments in filtration efficiency test. The plausible reason is that the electric charges of polypropylene filtration layer were neutralized after the treatment, especially with the use of organic solvents [13].

## V. Limitations

This study had only evaluated a surgical mask in the filtration of 0.1 – 1 µm NaCl droplets. However, the performance of the surgical mask after treatment has not been tested with more brands, particles of various sizes and different flowrates. As the study result was based on a modified protocol of NIOSH NaCl respirator certification test, it only provides comparisons among different groups of treatments. The study did not determine whether the treated samples could pass any international standards of surgical mask testing.

Future studies can focus on certain treatments, and investigate the optimal condition of decontamination treatment so as to minimize damage to the mask and to determine detailed disinfection conditions and kinetics that were required for other strains of microorganisms.

## VI. CONCLUSION

Decontamination and reuse of surgical mask become an alternative strategy to regenerate basic protective equipment so as to control the spread of disease under the current difficult situation. Non-fluid contacting disinfection methods such as UVC irradiation and dry heat retained the highest performance regarding filtration efficiency, structural consistence and surface hydrophobicity even after three cycles of treatments. These two disinfection methods for surgical mask would be considered under the severe PPE shortage situation.

## Data Availability

Authors confirm all supporting data are available within the article.

